# Chronic sensing during subcallosal cingulate deep brain stimulation captures ketamine-associated changes in major depressive disorder

**DOI:** 10.64898/2026.09.08.26362080

**Authors:** Srdjan Sumarac, Tristan Lawson, Leonardo Favi Bocca, Jennifer S Rabin, Peter Giacobbe, Sean Nestor, Clement Hamani, Nir Lipsman, Luka Milosevic, Benjamin Davidson

## Abstract

Ketamine and subcallosal cingulate (SCC) deep brain stimulation (DBS) have each shown antidepressant effects in treatment-resistant depression, yet how ketamine modulates SCC physiology during chronic stimulation remains poorly understood. We analyzed 5,907 hours of bilateral SCC sensing, 485 participant-triggered recordings across 193 days, 47 ketamine dosing episodes, and an in-clinic session. Across naturalistic and in-clinic recordings, ketamine was associated with a convergent bilateral shift toward 21–40 Hz fast-band activity. Post-dose snapshots showed lower aperiodic-adjusted 5–20 Hz power and higher 21–40 Hz power, while 23.44 Hz activity increased bilaterally after administration. Longitudinal 23.44 Hz tracking revealed larger bilateral increases around anxious and sad events, beginning before self-report and peaking approximately 15 minutes later, smaller decreases after neutral and happy events, and smaller lateralized changes around dosing, despite limited spectral separation between affective states at event onset. These findings provide direct human evidence that ketamine exposure and affective states can engage the same SCC frequency range with distinct spectral and temporal expressions. This positions SCC fast-band activity as a context-dependent signal whose interpretation may depend on its magnitude, laterality, and time course, motivating adaptive DBS strategies that decode evolving SCC dynamics rather than rely on simple band-power thresholds.

## Introduction

Major depressive disorder (MDD) is a leading cause of disability worldwide, and many patients do not achieve remission despite multiple pharmacologic and neuromodulatory treatments^1,2^. Ketamine and subcallosal cingulate (SCC) deep brain stimulation (DBS) represent distinct treatment approaches for individuals with treatment-resistant depression (TRD), although their physiological effects in humans remain incompletely understood^3,4^. Ketamine produces rapid, albeit often transient, reductions in depressive symptoms^3^, while SCC-DBS can yield sustained improvement in some patients with severe TRD^4^. Most human neurophysiological insight into ketamine has come from scalp electroencephalography (EEG), magnetoencephalography (MEG), or intracranial recordings in epilepsy^5^, leaving its effects on the deep limbic circuitry implicated in MDD poorly understood.

The SCC is a central node within the neurocircuitry of depression, serving as a convergence point for limbic, cognitive, and visceromotor networks and showing reproducible hyperactivity across imaging studies of MDD^6,7^. Modulation of this region through DBS can reorganize distributed affective circuits and produce antidepressant effects^4^. Sensing-enabled DBS systems permit chronic local field potential (LFP) recordings^8^, and recent SCC studies have identified cingulate dynamics associated with depression recovery during DBS^9^. Longitudinal SCC LFP recordings have further shown that 1/f activity varies with time of day and may track treatment response^10^. However, shorter-timescale SCC dynamics remain poorly characterized across naturalistic behavioral, temporal, stimulation, and pharmacologic contexts. In particular, how ketamine exposure is reflected in SCC physiology in vivo during ongoing DBS remains unknown.

To address this gap, we leveraged a densely sampled dataset of bilateral SCC recordings from a participant receiving SCC-DBS and repeated intranasal ketamine, spanning months of naturalistic home monitoring and an in-clinic ketamine session. Specifically, we (1) analyzed participant-triggered spectral snapshots to identify frequency-specific differences across affective states and ketamine exposure, (2) evaluated continuous SCC recordings acquired during the in-clinic ketamine session to characterize acute post-dose dynamics, and (3) examined longitudinal device bandpower to determine whether this fast-band activity showed context-specific temporal dynamics around self-reported affective and medication-related events.

## Methods

### Participant and clinical course

A woman with long-standing treatment-resistant major depressive disorder (TRD) participated in this study. Before enrollment, she had undergone multiple unsuccessful treatments, including antidepressant medications, psychotherapy, repetitive transcranial magnetic stimulation, and electroconvulsive therapy. She was receiving twice-weekly intranasal ketamine as part of her outpatient care, and this regimen remained unchanged throughout DBS evaluation, implantation, and follow-up. Ketamine was administered twice weekly using a 100 mg/mL formulation, with a total dose of 300 mg per administration. Except for the single in-clinic session described below, ketamine was self-administered at home. The study was approved by the Sunnybrook Research Institute Research Ethics Board (REB no. 277-2018), and the participant provided informed consent for the use and publication of her clinical data. The study was conducted in accordance with the Declaration of Helsinki. The work was conducted as a single-participant case study and was not registered as a clinical trial. Given the severity and chronicity of her symptoms, she underwent bilateral implantation of an SCC-DBS system (Medtronic Percept™ PC).

Stimulation was initially delivered in a monopolar configuration using a 1-minute-on/1-minute-off cycling pattern and was titrated from 2 to 6 mA at 125 Hz with a 60 μs pulse width. Cycling was discontinued before the Week 20 in-clinic session, and the stimulation amplitude was subsequently increased to 7 mA by Week 24. The pulse width was increased to 90 μs at Week 39, followed by further increases in stimulation amplitude to 7.5 mA by Week 44 and 8 mA by Week 54. Programming adjustments were made every two to four weeks. HAMD scores initially decreased from 21 at baseline to 11 at Week 16, followed by a transient increase to 16 at Week 17 and 20 at Week 20. Scores subsequently decreased to 13 at Week 24, remained relatively stable at 14 at Week 40, and improved further to 12 at Week 48 and 11 at Week 52. By Week 52, the HAMD score had decreased by 47.6% from baseline (21 to 11). Figure 1B summarizes the HAMD trajectory alongside the corresponding changes in stimulation parameters. The participant reported clinically meaningful symptomatic benefit from ketamine and adhered to the twice-weekly regimen throughout follow-up. The combined treatment course was tolerated without reported adverse or unanticipated events.

**Figure 1.**
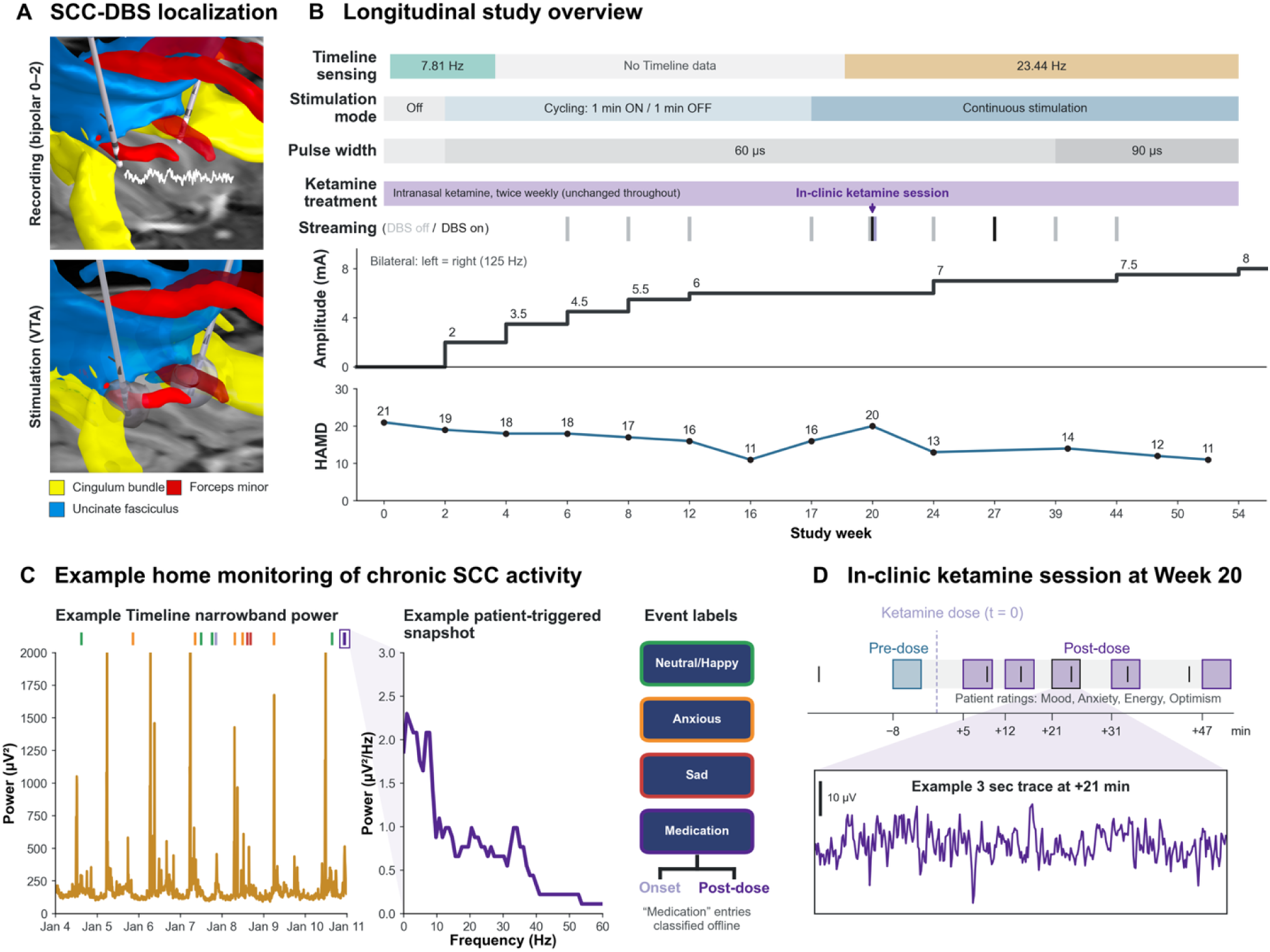
Study overview and chronic SCC sensing during ketamine treatment. (A) Three-dimensional reconstruction of bilateral SCC-DBS electrodes relative to the cingulum bundle (yellow), forceps minor (red), and uncinate fasciculus (blue). The upper reconstruction shows the bipolar 0–2 contact configuration used for SCC sensing, while the lower reconstruction shows the active stimulation contacts and simulated volumes of tissue activated (gray). (B) Longitudinal overview of stimulation mode and amplitude, pulse width, Timeline sensing configuration, intranasal ketamine treatment, and HAMD scores across follow-up. Timeline sensing was initially configured at 7.81 Hz and later at 23.44 Hz; the Week 20 in-clinic ketamine session is indicated. (C) Home monitoring included chronic Timeline narrowband power and participant-triggered 30-s spectral snapshots labeled Neutral/Happy, Anxious, Sad, or Medication. Medication entries were curated offline as Medication Onset or Medication Post-dose. Representative recordings are shown. (D) Week 20 in-clinic ketamine protocol showing one pre-dose and five post-dose SCC recordings over approximately 47 min, with a representative 3-s trace from +21 min. HAMD, Hamilton Depression Rating Scale.

### Surgical procedure and device configuration

Bilateral DBS electrodes (Model B33005, Medtronic Inc.) were implanted in the SCC white matter using MRI-guided stereotactic targeting under conscious sedation. Electrode trajectories were selected to traverse the cingulum bundle and approach the intersection of the forceps minor and uncinate fasciculus, consistent with established SCC targeting for depression^11–13^. Under general anesthesia, the electrodes were connected via extension cables to a Percept^™^ PC pulse generator implanted subcutaneously in the right chest. Right-sided pulse-generator placement was used to reduce electrocardiographic contamination risk^14^. Postoperative imaging confirmed electrode placement (Figure 1A).

After recovery, BrainSense^™^ was activated for chronic bilateral LFP recording using the bipolar 0–2 contact pair. The initial Timeline configuration tracked power centered at 7.81 Hz with a 5 Hz bandwidth. This was a clinical programming setting established before event-level spectra were available. After interim event spectra showed modulation in the high-beta range, sensing was reprogrammed to 23.44 Hz, the nearest available device center frequency. The 7.81 Hz Timeline dataset comprised 573 hours, including 240 hours with stimulation off and 333 hours during 2 mA stimulation delivered with a 1-minute-on/1-minute-off duty cycle; since Timeline stores 10-minute averages, these periods were recorded as approximately 0.9–1.0 mA. The 23.44 Hz Timeline dataset was acquired during continuous, noncycling DBS at 6–8 mA.

The patient controller provided four predefined event labels (Neutral/Happy, Anxious, Sad, and Medication). Each event entry triggered a 30-second bilateral LFP snapshot with an on-device power spectral density estimate. During home use, the participant was instructed to record mood-related events and to enter Medication at reported administration and approximately every 10 minutes for up to 60 minutes. Medication entries were subsequently curated offline by dosing sequence. The first entry was designated Medication Onset, and subsequent entries associated with that dosing sequence were designated Medication Post-dose. Entries were reviewed chronologically and grouped into distinct temporal clusters corresponding to individual dosing episodes. In the retained data, consecutive entries within a sequence were separated by no more than 86 minutes, whereas the next sequence began at least 70 hours later. All analyzed event snapshots were acquired with stimulation cycling disabled.

### Electrode localization

Lead localization and anatomical reconstruction were performed using Lead-DBS version 3.2^15^. Postoperative T1-weighted MRI was coregistered to the preoperative scan using SPM12 and normalized to MNI152 2009c space using the SyN algorithm in Advanced Normalization Tools (ANTs)^16^. Electrodes were manually localized on the postoperative scan by an expert operator (L.F.B.). The midcommissural point was identified using 3D Slicer version 5.9.0^17^, and ventral and dorsal contact coordinates were reported in millimeters relative to this native-space reference and after transformation to MNI152 nonlinear 2009c asymmetric standard space (Supplementary Table 1). Volumes of tissue activated (VTAs) were computed in native space using the volume-conductor model implemented in OSS-DBS^18^. Preoperative diffusion-weighted imaging (DWI) was preprocessed in FSL using topup and eddy current correction^19^, followed by automated fiber tracking and manual curation in DSI Studio (Hou version)^20^. Reconstructed fiber bundles were coregistered to the T1-weighted anatomy and visualized alongside the three-dimensional electrode reconstructions.

### Event-based snapshot analysis

Event-based spectral analysis included 485 bilateral snapshots collected across 193 local calendar days, comprising 168 Neutral/Happy, 95 Anxious, 70 Sad, 58 Medication Onset, and 94 Medication Post-dose events. Spectra were analyzed from 2–60 Hz across 59 frequency bins and parameterized using SpecParam with a fixed aperiodic model, peak widths of 3–12 Hz, a maximum of eight peaks, a minimum peak height of 0.1, and a peak threshold of 1.5^21^. Observed power was divided by the fitted aperiodic component to obtain an aperiodic-adjusted power ratio. Figures 2A and 2B show the mean raw and aperiodic-adjusted spectra, respectively, with 95% confidence intervals for each event category. Pointwise two-sided 95% confidence intervals were calculated across event snapshots at each frequency as the category mean plus or minus the Student’s t critical value multiplied by the sample standard error, with degrees of freedom equal to n minus 1.

**Figure 2.**
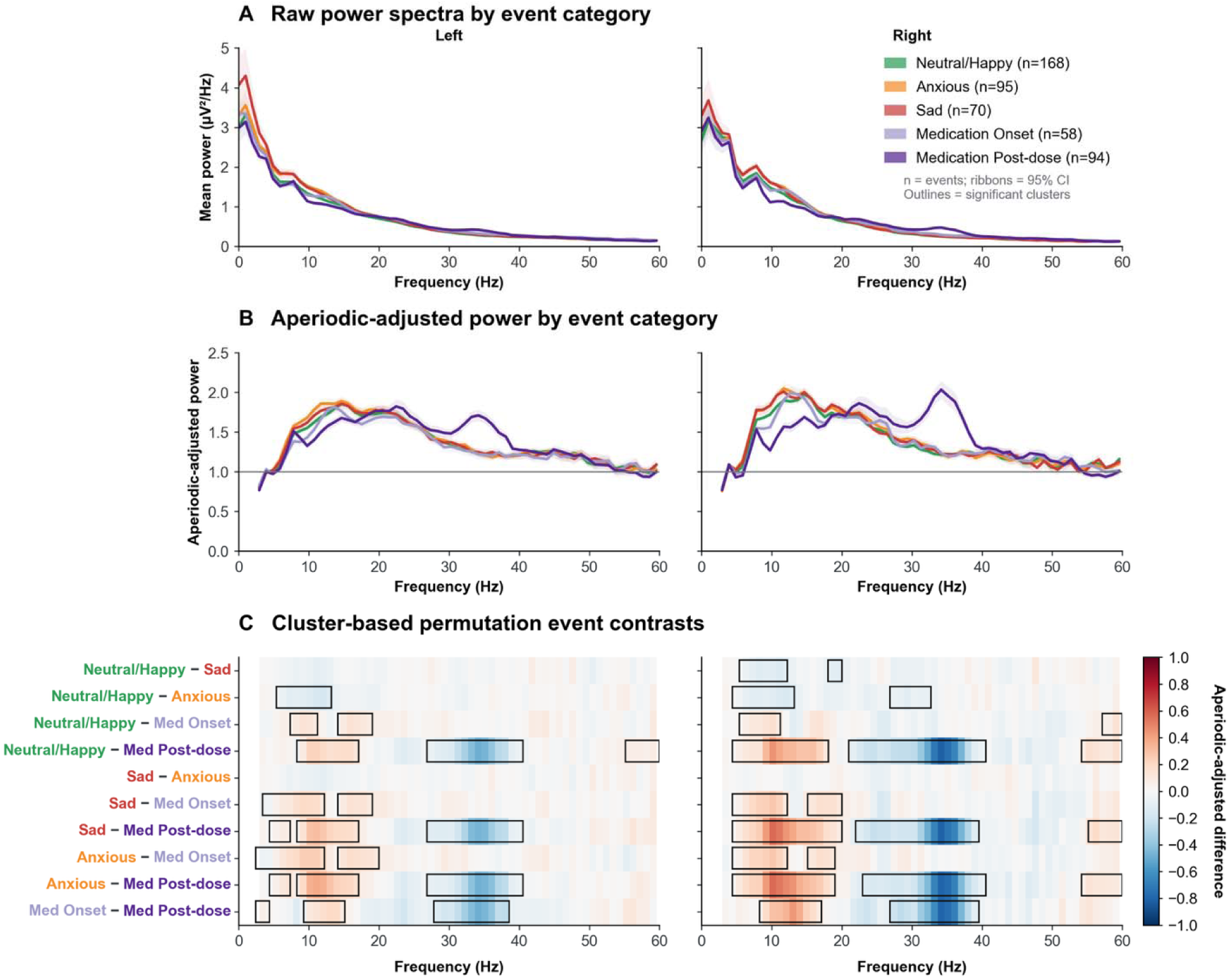
Spectral signatures of affective and medication-related events. (A) Mean raw power spectral density across event categories for the left and right SCC. Shading indicates 95% confidence intervals, and event counts are shown in the legend. (B) Mean aperiodic-adjusted power ratio. Shading indicates the 95% confidence interval, and the horizontal line denotes a ratio of 1. (C) Pairwise differences in aperiodic-adjusted power at each frequency, expressed as the first condition minus the second. Red indicates greater power in the first condition and blue greater power in the second. Black outlines denote significant clusters from cluster-based permutation testing with maximum-cluster correction across frequency. Complete cluster ranges, cluster masses, and permutation-corrected p values are reported in Supplementary Table 2. Covariate-adjusted 23.44 Hz sensitivity estimates are reported in Supplementary Table 4.

All 10 pairwise event-category contrasts were evaluated separately for each hemisphere, with individual snapshots treated as event-level observations. Differences were assessed at each frequency using a two-group one-way ANOVA F statistic, and adjacent bins meeting a p < .05 cluster-forming threshold were grouped. Cluster mass was calculated as the sum of F statistics across each cluster. Cluster significance was determined using 2,000 label configurations (the observed labels and 1,999 event-label permutations) and maximum-cluster correction across 2–60 Hz within each hemisphere and pairwise comparison. Clusters with permutation-corrected p ≤ .05 were considered significant. The F statistic is unsigned, so effect direction was determined from the observed signed mean difference between the first and second conditions, as displayed in Figure 2C. The primary naturalistic comparisons were unadjusted and interpreted as within-participant associations.

Sensitivity analyses evaluated 23.44-Hz aperiodic-adjusted power using separate hemisphere-specific ordinary least-squares models including event category, circular local clock time, nearest hemisphere-specific Timeline stimulation amplitude and pulse width within 10 minutes, and linear study date. Standard errors were clustered by local calendar day. Circular time was represented by paired sine and cosine terms. Benjamini–Hochberg false-discovery-rate correction was applied across eight planned contrasts. For the dosing-episode analysis, Post-dose spectra were averaged within each of 47 episodes and subtracted from the matched Medication Onset spectrum. Paired differences were tested using a two-sided cluster-based sign-flip test with 20,000 sign flips, a pointwise (p<.05) threshold, cluster mass defined as the sum of (|t|), and a single permutation maximum across hemispheres. Additional details are provided in Supplementary Tables 4 and 6 and Supplementary Fig. 3.

### In-clinic streaming session during Week 20

To characterize acute SCC dynamics around ketamine dosing, an in-clinic continuous recording session was performed during Week 20. Six bilateral BrainSense recordings from contact pair 0–2 were acquired during continuous DBS at 6 mA per hemisphere, 125 Hz, and 60 μs. One recording preceded dosing and five followed. Ketamine administration was annotated at approximately 7.46 p.m., with recording windows beginning approximately −8, +5, +12, +21, +31, and +47 minutes relative to dosing. Between recordings, the participant rated mood, anxiety, energy, and optimism while seated with eyes open. Ratings used a 0–9 numerical scale. Higher scores indicated better mood, greater anxiety, greater energy, and greater optimism, respectively.

Streaming signals were analyzed using time-frequency decomposition and Welch spectral estimation. Signals were sampled at 250 Hz. Spectrograms used 256-sample Tukey windows, 128-sample (50%) overlap, and a 512-point fast Fourier transform. The aperiodic-adjusted spectrograms were smoothed with a two-dimensional Gaussian kernel with σ = 1.0 frequency bin and 1.2 time bins. Welch spectra used 256-sample Hann windows with 50% overlap and 0.977 Hz frequency spacing. SpecParam was applied using the same peak settings as the event analysis, with models fit over 1–80 Hz for the spectrograms and 2–60 Hz for the power spectra. Power was divided by the fitted aperiodic component before analysis. Given the single-session design, changes at 7.81 and 23.44 Hz were expressed relative to the pre-dose baseline and examined alongside the clinical ratings without inferential testing. Complete bilateral raw traces from all six recordings were visually reviewed for stereotyped rhythmic, bilaterally synchronous sharp deflections consistent with electrocardiographic contamination.

### Longitudinal timeline and event-day analysis

The longitudinal dataset comprised 573 hours recorded at 7.81 Hz and 5,334 hours recorded at 23.44 Hz. Timeline power was z-normalized separately at each sensing frequency using the mean and sample standard deviation of pooled left- and right-hemisphere observations from the corresponding recording period. Neutral/Happy, Anxious, Sad, and Medication Onset events were examined. Medication Post-dose events were not included because multiple post-dose entries could arise from a single dosing sequence.

For each event, Timeline observations from −100 to +100 minutes were averaged into 10-minute bins. Each trajectory was referenced to the mean of available 10-minute bins from the event-free −200 to −100 minute baseline. A target event was excluded if any curated event occurred within its baseline interval. Repeated events of the same category on the same local calendar date were averaged so that the event-day was the sampling unit. Only event-day trajectories containing data in all 20 event-window bins were included in inferential testing.

At each time bin, baseline-corrected power was compared with zero using a two-sided one-sample t test. Adjacent bins exceeding an uncorrected cluster-forming threshold of p < .05 were grouped, and cluster mass was calculated as the sum of squared t statistics across each cluster. Complete event-day trajectories were sign-flipped, preserving their temporal dependence. Cluster-corrected p values were obtained from the maximum cluster mass across the −100 to +100 minute window. All possible sign-flip configurations were evaluated for analyses with 10 or fewer complete event-days, whereas 2,000 configurations (the observed configuration plus 1,999 pseudorandom sign flips) were evaluated for analyses with 11 or more complete event-days. Analyses were performed separately for each event category, sensing frequency, and hemisphere, and clusters with corrected p ≤ .05 were considered significant.

Figure 4C shows the mean and pointwise 95% confidence interval across complete event-days, while Figure 4D summarizes the corresponding mean event-minus-baseline changes and significant clusters. Pointwise two-sided 95% confidence intervals were calculated across complete event-day trajectories at each 10-minute bin as the mean plus or minus the Student’s t critical value multiplied by the event-day standard error, with degrees of freedom equal to the number of complete event-days minus 1. Timeline contains device-defined narrowband estimates rather than full spectra, so periodic and aperiodic components could not be separated. Z-scored Timeline power was also summarized descriptively by local clock time on ketamine treatment dates and the following calendar dates (Supplementary Fig. 1). An isolated-event sensitivity analysis repeated the analysis after excluding target events with any other curated event within −100 to +100 minutes (Supplementary Table 5). Generative artificial intelligence (AI) tools were used to assist with research and manuscript preparation, with all outputs reviewed and verified by the authors.

## Results

### Contact localization

Bilateral electrodes were localized within the SCC white matter, with the bipolar 0–2 recording configuration and stimulation VTAs shown relative to the forceps minor, uncinate fasciculus, and cingulum bundle (Figure 1A). Recording contact coordinates are provided in Supplementary Table 1. The simulated VTAs overlapped the targeted fiber pathways, including pathways previously associated with therapeutic SCC-DBS^11–13,22^.

### Event-related spectral differences and ketamine-associated changes

Aperiodic-adjusted spectra showed a bilateral reorganization during Medication Post-dose events, with lower power at lower frequencies and higher power across the beta-gamma range relative to all other event categories (Figure 2B, C). Event timing differed across categories, with Neutral/Happy events occurring predominantly in the evening, Anxious and Sad events earlier in the day, and Medication events primarily at night. Complete frequency ranges, cluster masses, and permutation-corrected p values are reported in Supplementary Table 2. Effect direction is shown in Figure 2C. Although the post-dose spectral reorganization was bilateral, its frequency extent was asymmetric. Across Post-dose contrasts, higher-power clusters began at lower frequencies and lower-power clusters extended to higher frequencies on the right than on the left. At 23.44 Hz, Post-dose power was significantly higher than each comparison category in both hemispheres after adjustment for clock time, stimulation amplitude, pulse width, and study date (all BH-FDR q ≤ .0136; Supplementary Table 4). The bilateral fast-band difference also remained significant when Post-dose snapshots were averaged within 47 dosing episodes and compared with matched onsets (both p < .0001; Supplementary Fig. 3 and Supplementary Table 6).

### In-clinic ketamine response dynamics

During the Week 20 in-clinic session, aperiodic-adjusted 23.44 Hz power increased after ketamine administration in both hemispheres, with a larger increase in the right SCC (Figure 3). The increase emerged during the early post-dose recordings and persisted through the final recording. At 7.81 Hz, adjusted power increased above baseline on the left but remained below baseline on the right. Mood and optimism increased during the early post-dose period, anxiety initially decreased, and energy increased transiently before falling below baseline. The acute 23.44 Hz response was bilateral but asymmetric in magnitude, whereas the 7.81 Hz response differed in direction between hemispheres.

**Figure 3.**
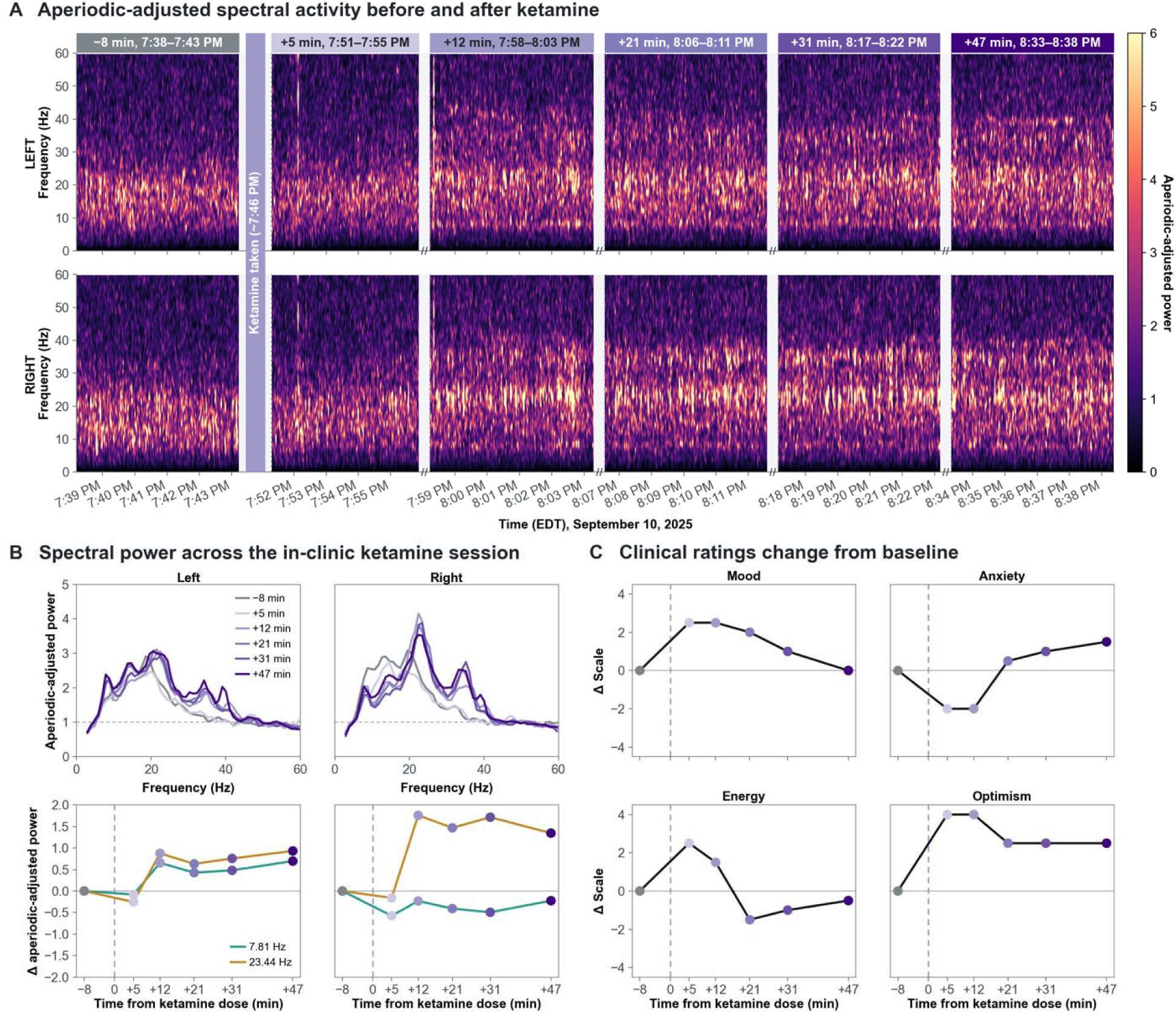
Aperiodic-adjusted SCC activity during the Week 20 in-clinic ketamine session. (A) Bilateral contact 0–2 spectrograms from one pre-dose and five post-dose BrainSense recordings acquired during continuous DBS at 6 mA per hemisphere. Power was divided by the fitted aperiodic component. The vertical purple band marks ketamine administration at approximately 7:46 p.m. Post-dose recordings began approximately +5, +12, +21, +31, and +47 min after dosing. (B) Aperiodic-adjusted Welch power spectra for the left and right SCC across all six recordings (top), with changes from pre-dose baseline at 7.81 and 23.44 Hz plotted against time from ketamine administration (bottom). (C) Changes from pre-dose baseline in mood, anxiety, energy, and optimism ratings across the session. Vertical dashed lines indicate ketamine administration at 0 min.

### Longitudinal power and event-linked dynamics

We next examined how SCC narrowband activity evolved longitudinally around affective and medication-related events. The longitudinal dataset comprised 573 hours at 7.81 Hz and 5,334 hours at 23.44 Hz (Figure 4A). No event-related clusters at 7.81 Hz survived cluster-level correction. At 23.44 Hz, Neutral/Happy events were followed by small bilateral decreases, extending from +10 to +100 minutes on the left (mean Δz = −0.14, permutation-corrected p = .0005) and +50 to +100 minutes on the right (mean Δz = −0.11, p = .0025). In contrast, Anxious events showed larger bilateral increases beginning before the logged event and peaking at +15 minutes, with principal clusters from −20 to +100 minutes on the left (mean Δz = 0.89, p = .0005) and −20 to +50 minutes on the right (mean Δz = 0.96, p = .0005). Sad events showed a similar pattern, with increases from −20 to +60 minutes on the left (mean Δz = 0.85, p = .0005) and −20 to +30 minutes on the right (mean Δz = 1.03, p = .0005), also peaking at +15 minutes. Medication Onset showed smaller, lateralized changes. Power decreased from +80 to +100 minutes on the left (mean Δz = −0.10, p = .023) and from −40 to −20 minutes on the right (mean Δz = −0.14, p = .0205), then increased from +10 to +40 minutes on the right (mean Δz = 0.33, p = .027). Additional smaller clusters were observed outside the principal event-related windows and are reported in Supplementary Table 3. Complete cluster ranges, cluster masses, and permutation-corrected p values are reported in Supplementary Table 3. Overall, the longitudinal effects were broadly bilateral but differed in magnitude and duration between hemispheres, while Medication Onset changes were smaller and more lateralized. The principal 23.44 Hz affective-event clusters remained significant after excluding overlapping events (all corrected p ≤ .015; Supplementary Table 5).

**Figure 4.**
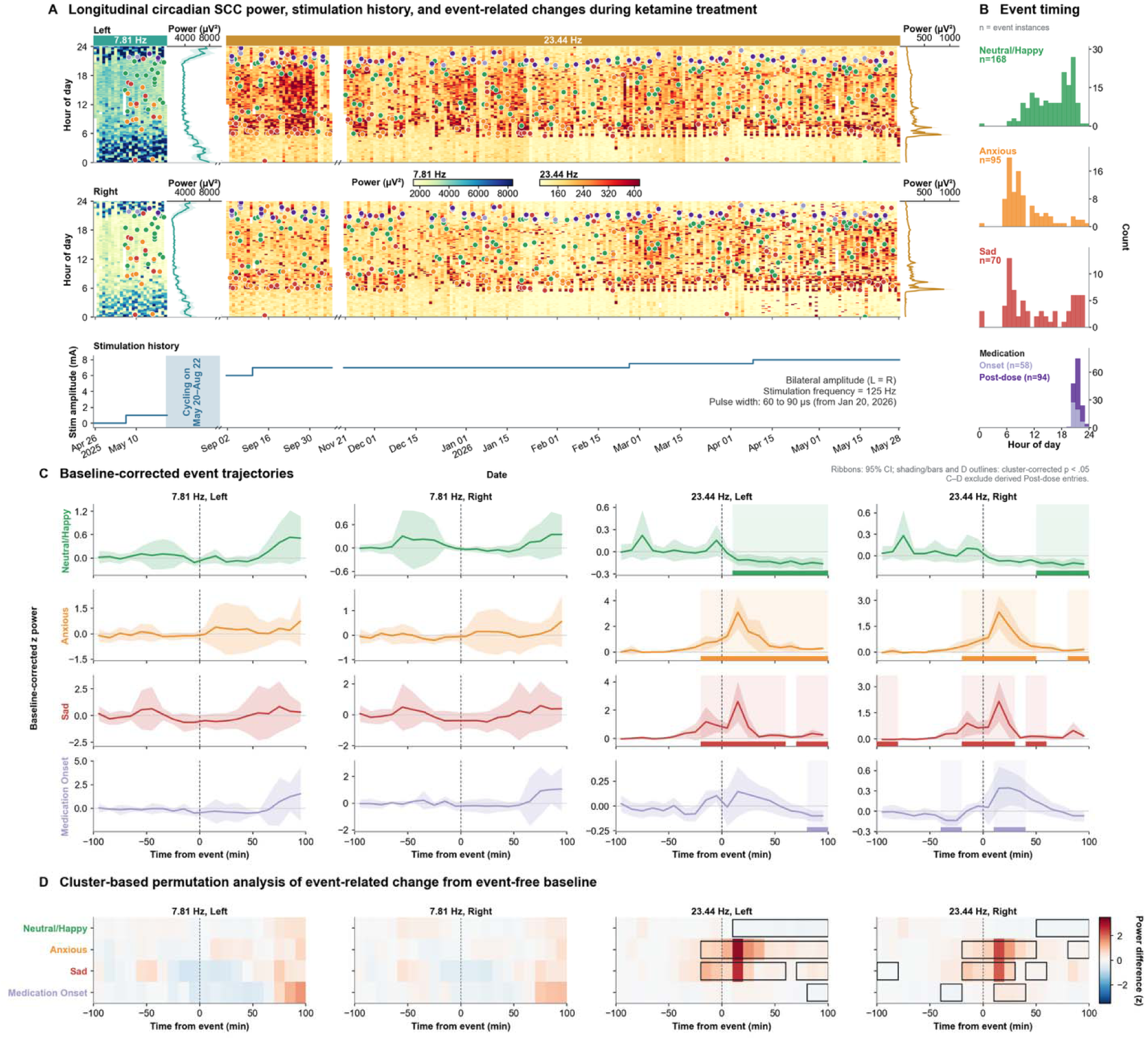
Longitudinal SCC power, stimulation history, and event-linked trajectories. (A) Clock-time heatmaps of bilateral SCC power at 7.81 and 23.44 Hz across the longitudinal recording period, with event markers, mean clock-time power profiles, and stimulation history. (B) Clock-time distributions of included events. Medication Onset and Medication Post-dose are shown separately. (C) Baseline-corrected event-day trajectories from −100 to +100 min for Neutral/Happy, Anxious, Sad, and Medication Onset events. Medication Post-dose entries were excluded because multiple entries could arise from a single dosing sequence. Lines indicate means and shading 95% confidence intervals across event-days; colored strips denote significant temporal clusters from cluster-based permutation testing. (D) Mean event-minus-baseline change across relative time. Black outlines denote significant clusters identified by cluster-based permutation testing with maximum-cluster correction across the −100 to +100 min window. Complete cluster statistics are reported in Supplementary Table 3, with the isolated-event sensitivity analysis in Supplementary Table 5.

## Discussion

Chronic SCC sensing identified convergent spectral and temporal changes associated with ketamine exposure and self-reported affective state during ongoing DBS. In patient-triggered recordings collected during home monitoring, Medication Post-dose events were characterized by a bilateral redistribution of aperiodic-adjusted power from approximately 5–20 Hz toward 21–40 Hz. The in-clinic streaming session showed a similar bilateral increase across the same fast-band range following ketamine administration. Within this range, 23.44 Hz was the device-defined center frequency selected for chronic sensing, allowing us to examine its longitudinal dynamics around medication-related and affective events. At 23.44 Hz, Anxious and Sad events were associated with bilateral increases beginning before self-report and peaking approximately 15 minutes later, whereas Neutral and Happy events were followed by smaller decreases. Medication Onset produced smaller, more lateralized changes. Although these longitudinal effects were broadly bilateral, their magnitude and duration differed between hemispheres. The ketamine and affective-state findings identify a patient-specific fast-band physiological feature that is strongly modulated by ketamine and shows distinct dynamics during negative affect. At 23.44 Hz, affective state was reflected less by power in a single event snapshot than by its trajectory over tens of minutes. These findings position SCC fast-band activity as a context-dependent signal whose magnitude, laterality, and temporal trajectory vary across ketamine exposure and negative affect.

### Interpretation of SCC spectral changes in the context of ketamine

Multiple complementary pathways have been proposed to explain ketamine’s rapid antidepressant effects, including circuit-level disinhibition, synaptic plasticity, and metabolic signaling^23,24^. Ketamine can increase cortical excitability through NMDA receptor blockade on inhibitory interneurons and engage AMPA receptors, BDNF signaling, and downstream plasticity pathways over longer timescales^23,25^. Opioid signaling may also contribute to ketamine’s antidepressant effects, with human evidence implicating μ-opioid mechanisms^26^ and preclinical work implicating β-endorphin signaling^27^. The spectral pattern observed in the SCC was broadly consistent with previous human electrophysiological studies reporting increased beta and gamma activity and reduced alpha or theta power after ketamine^5,28–31^. To separate frequency-specific activity from the aperiodic 1/f background, the event-based and in-clinic spectra were analyzed after removal of the fitted aperiodic component^21^. Medication Post-dose was characterized by reduced lower-frequency power and increased power across approximately 21–40 Hz, demonstrating a frequency-specific SCC spectral shift associated with ketamine exposure. The ketamine-associated shift overlapped the fast-band range in which 23.44 Hz activity evolved around Anxious and Sad events. Ketamine robustly engaged this frequency range but produced a distinct bilateral spectral shift. The shared frequency range but divergent spectral and temporal expression suggest that SCC fast-band activity is strongly dependent on physiological context. These recordings define an electrophysiological response at the SCC, while future mechanistic studies can resolve the molecular pathways that produce it. The time course of the spectral changes may also help contextualize neuroimaging studies reporting both increased and decreased SCC activity after ketamine^29,32–34^, as differences in acquisition method and sampling time may capture different phases of the response. Preclinical studies support a distributed network interpretation, with ketamine suppressing NMDA receptor-dependent bursting in the lateral habenula and altering low-frequency activity associated with negative behavioral states^35^. SCC-DBS has also been associated with changes in habenular-frontocingulate connectivity^36^, while SCC connectivity with dorsolateral prefrontal cortex relates to antidepressant response to transcranial magnetic stimulation^37^. Collectively, these observations place the observed SCC response within a distributed cingulate-prefrontal-habenular network.

### Temporal context and event interpretation

Longitudinal SCC activity showed distinct temporal patterns across event categories. Anxious and Sad events shared similar 23.44 Hz trajectories, with bilateral increases beginning before self-report and peaking approximately 15 minutes later despite no spectral separation at the logged event. Neutral/Happy events were followed by smaller decreases, whereas Medication Onset showed smaller, lateralized deviations. The fast-band trajectory, rather than a static event-locked spectrum, revealed diverging dynamics across negative-affect and Neutral/Happy events. SCC bandpower also varied systematically across the day, and event categories showed distinct clock-time distributions, with Anxious and Sad events concentrated earlier in the day, Neutral/Happy events predominantly in the evening, and Medication events at night. This pattern is consistent with prior chronic SCC recordings showing that electrophysiological features vary with time of day^10^. Accordingly, the observed SCC signal should be interpreted in relation to both event-related dynamics and broader time-of-day variation. The event-specific baseline used for the longitudinal analysis quantifies changes relative to the local physiological state preceding each event, and the affective-event patterns persisted after exclusion of overlapping events. Supplementary Fig. 1 provides additional descriptive context for within-day variation in SCC power on medication days and the following calendar days. The affective-event patterns persisted after exclusion of overlapping events.

### Implications for chronic sensing and adaptive DBS

Prior SCC studies have identified longitudinal 1/f activity^10^ and a multivariate cingulate-recovery signal^9^ as markers of longer-term change during DBS. The present findings extend this work to minute-scale dynamics by showing that ketamine and negative affect engage a shared fast-band range but with different spectral and temporal expressions. Ketamine produced a robust bilateral shift toward 21–40 Hz activity, whereas negative affect was reflected in the evolving magnitude and laterality of 23.44 Hz power over tens of minutes. Thus, the same local frequency feature may carry different meanings depending on its trajectory and physiological context. Consistent with evidence that stimulation effects depend on baseline state and that patient-specific neural responses can guide programming or trigger stimulation^38–40^, these findings suggest that adaptive DBS should infer an evolving circuit state rather than respond to a single high or low band-power value. Electrophysiological studies in Parkinson’s disease provide a precedent for extending this decoding framework beyond local power. Therapeutic DBS has been shown to modulate cortical phase–amplitude coupling^41^, motor-cortical activity has been used to control adaptive stimulation^42^, and chronic recordings across cortex and basal ganglia can distinguish clinically relevant motor states^43^. Analogously, simultaneous SCC and cortical or other network-level recordings could determine whether similar SCC fast-band changes reflect distinct affective or treatment states depending on their interregional coherence or cross-frequency coupling. Multisite sensing could add a complementary spatial dimension, as activity may propagate through a target without corresponding changes in local signal magnitude^44^, while cortical biomarkers may capture information unavailable from the DBS lead alone^45^. Integrating signal trajectories, within-target spatial patterns, and distributed network coupling therefore offers a concrete path toward more specific state inference for adaptive DBS, although its generalizability must be tested across participants and repeated dosing.

### Limitations and future directions

This report describes chronic SCC sensing in one participant with treatment-resistant depression receiving SCC-DBS and repeated ketamine therapy. The extensive within-participant sampling provides detailed physiological characterization across months of naturalistic recording, although generalizability to other patients remains limited. The effects were broadly bilateral but asymmetric across analyses. Potential contributors include small differences in lead or contact location relative to the targeted white-matter pathways, stimulation-field geometry and volume of tissue activated, electrode–tissue interface properties, underlying hemispheric anatomy or network organization, and residual recording or stimulation-related artifact. These possibilities cannot be distinguished in a single participant. Home recordings preserved real-world behavioral context but offered less experimental control: affective and medication-related events were unevenly distributed across clock time, and home ketamine dosing was identified from patient-entered event markers rather than independently synchronized timestamps. The Medication Post-dose category grouped repeated patient-entered markers following Medication Onset within home dosing sequences. These entries represent self-reported post-dose observations rather than fixed pharmacokinetic time points. The two Timeline frequencies were also recorded during different periods of the study and under different stimulation conditions, with 23.44 Hz selected after the event spectra indicated prominent fast-band modulation. The in-clinic findings came from one ketamine session and require replication across repeated sessions. A dedicated ECG channel was not recorded, so subtle cardiac contamination remains possible. The supplementary DBS-off/on comparison was unmatched and nonrandomized, and affective state was not standardized across recordings, so differences may reflect stimulation, concurrent state, or both. Prospective studies across repeated ketamine sessions under fixed sensing and stimulation conditions are needed to establish reproducibility and better separate the ketamine-related response from temporally distinct changes around spontaneous affective states. At 23.44 Hz, Post-dose power remained higher than all comparison categories in covariate-adjusted sensitivity models (all BH-FDR q ≤ .0136), but residual confounding by unmeasured behavioral or treatment factors remains possible.

## Conclusion

Chronic SCC sensing revealed a patient-specific fast-band feature engaged during both ketamine exposure and the temporal dynamics of negative affect. Ketamine exposure produced a bilateral, frequency-specific shift toward 21–40 Hz activity, while 23.44 Hz power evolved over tens of minutes around Anxious and Sad states despite limited separation at this frequency in the event snapshots. This overlap reveals a context-dependent SCC physiological feature whose expression differs across pharmacological and affective states. These findings motivate adaptive DBS strategies that decode evolving circuit dynamics rather than respond to a static spectral threshold. This framework can now be tested across patients and repeated dosing.

## Funding

The authors received no specific funding for this work.

## Author contributions

S.S. led the study design, data curation, analysis, visualization, interpretation, and manuscript preparation. T.L. contributed to data acquisition and initial manuscript preparation. L.F.B. contributed to data acquisition, imaging analyses and lead reconstruction, and reviewed the manuscript. J.S.R., P.G., S.N., C.H., and N.L. contributed to interpretation of the findings and reviewed the manuscript. L.M. and B.D. jointly supervised the work and contributed to study design and interpretation. All authors reviewed and approved the final manuscript.

## Competing interests

The authors declare no competing interests.

## Data availability

The de-identified data supporting the findings of this study are available from the corresponding author upon reasonable request.

## Code availability

Custom code supporting the analyses reported in this study is available from the corresponding author upon request.

## Notes

### Competing Interest Statement

The authors have declared no competing interest.

### Author Declarations

The Research Ethics Board of Sunnybrook Research Institute gave ethical approval for this work (REB no. 277-2018).

