## Supplementary Information for "Chronic sensing during subcallosal cingulate deep brain stimulation captures ketamine-associated changes in major depressive disorder"

**Supplementary Table 1.** **DBS lead-contact localization.** Ventral and dorsal contact coordinates are reported for each hemisphere in millimeters relative to the midcommissural point (MCP) and in MNI152 nonlinear 2009c asymmetric standard space. Coordinates are ordered x, y, z; negative and positive values denote left/right, posterior/anterior, and inferior/superior directions, respectively.

| **Side** | **Contact** | **Relative to MCP (x, y, z), mm** | **MNI152 2009c (x, y, z), mm** |
| --- | --- | --- | --- |
| Left | Ventral | (−9.9, 37.7, −6.1) | (−8.7, 27.6, −14.3) |
| Left | Dorsal | (−11.0, 37.3, −2.3) | (−10.5, 27.8, −9.4) |
| Right | Ventral | (5.1, 36.7, −5.6) | (9.5, 26.4, −13.7) |
| Right | Dorsal | (7.0, 36.9, −2.1) | (10.6, 26.7, −8.7) |

**Supplementary Table 2. Significant frequency-domain clusters for event-related spectral contrasts.** Significant clusters in aperiodic-adjusted spectra are reported separately by hemisphere for all 10 pairwise event-category comparisons. Only clusters surviving correction are listed; comparisons not shown had no significant clusters. Multiple rows for the same comparison represent separate, noncontiguous frequency clusters. Event counts were Neutral/Happy, n = 168; Anxious, n = 95; Sad, n = 70; Medication Onset, n = 58; and Medication Post-dose, n = 94. Effect direction is shown in the corresponding spectral contrasts in Figure 2C.

| **Hemisphere** | **Comparison** | **Cluster range** | **Cluster mass (sum F)** | **Corrected p value** |
| --- | --- | --- | --- | --- |
| Left | Anxious vs Medication Onset | 2.93 to 11.72 Hz | 223.204 | .0005 |
| Left | Anxious vs Medication Post-dose | 8.79 to 16.60 Hz | 508.688 | .0005 |
| Left | Anxious vs Medication Post-dose | 27.34 to 40.04 Hz | 681.016 | .0005 |
| Left | Medication Onset vs Medication Post-dose | 28.32 to 38.09 Hz | 497.522 | .0005 |
| Left | Neutral/Happy vs Medication Post-dose | 8.79 to 16.60 Hz | 261.294 | .0005 |
| Left | Neutral/Happy vs Medication Post-dose | 27.34 to 40.04 Hz | 991.944 | .0005 |
| Left | Sad vs Medication Post-dose | 8.79 to 16.60 Hz | 301.31 | .0005 |
| Left | Sad vs Medication Post-dose | 27.34 to 40.04 Hz | 532.116 | .0005 |
| Right | Anxious vs Medication Onset | 4.88 to 11.72 Hz | 188.556 | .0005 |
| Right | Anxious vs Medication Post-dose | 4.88 to 18.55 Hz | 1182.453 | .0005 |
| Right | Anxious vs Medication Post-dose | 23.44 to 40.04 Hz | 920.2 | .0005 |
| Right | Medication Onset vs Medication Post-dose | 8.79 to 16.60 Hz | 378.307 | .0005 |
| Right | Medication Onset vs Medication Post-dose | 27.34 to 39.06 Hz | 629.856 | .0005 |
| Right | Neutral/Happy vs Anxious | 4.88 to 12.70 Hz | 117.454 | .0005 |
| Right | Neutral/Happy vs Medication Post-dose | 4.88 to 17.58 Hz | 1103.648 | .0005 |
| Right | Neutral/Happy vs Medication Post-dose | 21.48 to 40.04 Hz | 1794.248 | .0005 |
| Right | Neutral/Happy vs Medication Post-dose | 54.69 to 59.57 Hz | 141.726 | .0005 |
| Right | Sad vs Medication Onset | 4.88 to 11.72 Hz | 201.003 | .0005 |
| Right | Sad vs Medication Post-dose | 4.88 to 19.53 Hz | 1269.022 | .0005 |
| Right | Sad vs Medication Post-dose | 22.46 to 39.06 Hz | 785.178 | .0005 |
| Left | Medication Onset vs Medication Post-dose | 9.77 to 14.65 Hz | 100.371 | .001 |
| Left | Neutral/Happy vs Anxious | 5.86 to 12.70 Hz | 109.653 | .001 |
| Left | Sad vs Medication Onset | 3.91 to 11.72 Hz | 116.006 | .001 |
| Right | Neutral/Happy vs Medication Onset | 5.86 to 10.74 Hz | 83.524 | .001 |
| Right | Neutral/Happy vs Sad | 5.86 to 11.72 Hz | 81.302 | .001 |
| Right | Sad vs Medication Post-dose | 55.66 to 59.57 Hz | 106.412 | .002 |
| Right | Anxious vs Medication Post-dose | 54.69 to 59.57 Hz | 84.696 | .0025 |
| Right | Sad vs Medication Onset | 15.63 to 19.53 Hz | 49.127 | .0025 |
| Left | Anxious vs Medication Onset | 14.65 to 19.53 Hz | 62.741 | .004 |
| Left | Anxious vs Medication Post-dose | 4.88 to 6.84 Hz | 60.84 | .004 |
| Right | Neutral/Happy vs Anxious | 27.34 to 32.23 Hz | 36.562 | .01 |
| Left | Sad vs Medication Onset | 14.65 to 18.55 Hz | 35.276 | .015 |
| Left | Neutral/Happy vs Medication Onset | 7.81 to 10.74 Hz | 37.16 | .019 |
| Left | Neutral/Happy vs Medication Onset | 14.65 to 18.55 Hz | 36.587 | .0195 |
| Right | Anxious vs Medication Onset | 15.63 to 18.55 Hz | 32.763 | .023 |
| Left | Sad vs Medication Post-dose | 4.88 to 6.84 Hz | 31.776 | .025 |
| Left | Neutral/Happy vs Medication Post-dose | 55.66 to 59.57 Hz | 30.067 | .0265 |
| Right | Neutral/Happy vs Sad | 18.55 to 19.53 Hz | 22.412 | .03 |
| Right | Neutral/Happy vs Medication Onset | 57.62 to 59.57 Hz | 22.668 | .038 |
| Left | Medication Onset vs Medication Post-dose | 2.93 to 3.91 Hz | 26.002 | .0415 |

**Supplementary Table 3. Significant temporal clusters of event-related SCC power.** Significant clusters of event-minus-baseline SCC power are reported separately by hemisphere for Neutral/Happy, Anxious, Sad, and Medication Onset. Medication Post-dose entries were excluded because multiple entries could arise from one dosing sequence. Only clusters surviving correction are listed; no significant clusters were identified at 7.81 Hz. Multiple rows for the same event represent separate temporal clusters. Effect direction is shown by the corresponding trajectories in Figure 4. The complete event-day count used for inference is reported for each cluster.

| **Hemisphere** | **Frequency** | **Event (vs baseline)** | **Complete event-days, n** | **Cluster range** | **Cluster mass (sum t²)** | **Corrected p value** |
| --- | --- | --- | --- | --- | --- | --- |
| Left | 23.44 Hz | Anxious | 72 | −20 to 100 min | 181.651 | .0005 |
| Left | 23.44 Hz | Neutral/Happy | 115 | 10 to 100 min | 135.91 | .0005 |
| Left | 23.44 Hz | Sad | 53 | −20 to 60 min | 78.039 | .0005 |
| Right | 23.44 Hz | Anxious | 72 | −20 to 50 min | 110.531 | .0005 |
| Right | 23.44 Hz | Sad | 53 | −20 to 30 min | 47.796 | .0005 |
| Right | 23.44 Hz | Neutral/Happy | 115 | 50 to 100 min | 64.309 | .0025 |
| Left | 23.44 Hz | Sad | 53 | 70 to 100 min | 21.264 | .0155 |
| Right | 23.44 Hz | Medication Onset | 46 | −40 to −20 min | 24.311 | .0205 |
| Left | 23.44 Hz | Medication Onset | 46 | 80 to 100 min | 27.035 | .023 |
| Right | 23.44 Hz | Anxious | 72 | 80 to 100 min | 12.208 | .0265 |
| Right | 23.44 Hz | Medication Onset | 46 | 10 to 40 min | 19.235 | .027 |
| Right | 23.44 Hz | Sad | 53 | 40 to 60 min | 13.876 | .027 |
| Right | 23.44 Hz | Sad | 53 | −100 to −80 min | 11.661 | .0395 |

Supplementary methods for covariate-adjusted sensitivity analysis. At 23.44 Hz, hemisphere-specific ordinary least-squares models included event category, local clock time as sine and cosine terms, the nearest hemisphere-specific Timeline stimulation amplitude and pulse width within 10 minutes, and linear study date. Standard errors were clustered by local calendar day, and the eight planned Post-dose contrasts were adjusted using the Benjamini–Hochberg false-discovery rate.

**Supplementary Table 4. Covariate-adjusted sensitivity analysis of 23.44 Hz event-snapshot power.** Adjusted Medication Post-dose differences are from the models described above. Positive differences indicate higher aperiodic-adjusted power during Medication Post-dose events. All 485 snapshots across 193 local calendar days were included.

| **Hemisphere** | **Comparison** | **Adjusted difference at 23.44 Hz (95% CI)** | **BH-FDR q value** |
| --- | --- | --- | --- |
| Left | Post-dose vs Neutral/Happy | .197 (.080 to .315) | .0044 |
| Left | Post-dose vs Anxious | .217 (.097 to .338) | .0037 |
| Left | Post-dose vs Sad | .178 (.053 to .303) | .0088 |
| Left | Post-dose vs Medication Onset | .187 (.062 to .311) | .0068 |
| Right | Post-dose vs Neutral/Happy | .225 (.075 to .375) | .0068 |
| Right | Post-dose vs Anxious | .191 (.047 to .336) | .0109 |
| Right | Post-dose vs Sad | .200 (.052 to .348) | .0109 |
| Right | Post-dose vs Medication Onset | .198 (.041 to .355) | .0136 |

**Supplementary Table 5. Isolated-event sensitivity analysis of longitudinal SCC power.** Target events were retained only when no other curated event, including Medication Post-dose, occurred within −100 to +100 minutes. The primary event-free −200 to −100-minute baseline exclusion, coverage requirements, and cluster-permutation settings were unchanged. Before coverage and baseline filtering, 127/168 Neutral/Happy, 78/95 Anxious, 61/70 Sad, and 4/58 Medication Onset markers were retained. Only significant 23.44 Hz clusters are listed, with the complete event-day count used for inference reported for each cluster. No Medication Onset event-days remained eligible for 23.44 Hz inference after coverage and baseline filtering.

| **Hemisphere** | **Frequency** | **Event (vs baseline)** | **Complete event-days, n** | **Cluster range** | **Cluster mass (sum t²)** | **Cluster-corrected p value** |
| --- | --- | --- | --- | --- | --- | --- |
| Left | 23.44 Hz | Anxious | 62 | −20 to 30 min | 87.872 | .0005 |
| Left | 23.44 Hz | Anxious | 62 | 40 to 100 min | 87.972 | .0005 |
| Left | 23.44 Hz | Sad | 47 | −20 to 60 min | 64.413 | .0005 |
| Right | 23.44 Hz | Anxious | 62 | −10 to 50 min | 99.263 | .0005 |
| Left | 23.44 Hz | Neutral/Happy | 94 | 10 to 100 min | 106.303 | .001 |
| Right | 23.44 Hz | Sad | 47 | −20 to 30 min | 41.443 | .001 |
| Right | 23.44 Hz | Neutral/Happy | 94 | 50 to 100 min | 60.847 | .003 |
| Left | 23.44 Hz | Sad | 47 | 70 to 100 min | 18.497 | .015 |

**Supplementary Table 6. Dosing-episode sensitivity analysis of Medication Post-dose spectral power.** Each Post-dose snapshot was assigned to its immediately preceding Medication Onset, aperiodic-adjusted Post-dose spectra were averaged within each dosing episode, and the matched onset spectrum was subtracted. Paired differences from 47 dosing episodes (94 Post-dose snapshots) were tested using 20,000 sign flips and a two-sided p < .05 cluster-forming threshold, with one permutation maximum across both hemispheres. Only globally significant clusters are listed; effect direction is shown in Supplementary Fig. 3.

| **Hemisphere** | **Comparison** | **Cluster frequency range** | **Cluster mass (sum \|t\|)** | **Globally corrected p value** |
| --- | --- | --- | --- | --- |
| Left | Post-dose vs Onset | 27.34 to 38.09 Hz | 70.640 | <.0001 |
| Right | Post-dose vs Onset | 27.34 to 39.06 Hz | 84.733 | <.0001 |
| Right | Post-dose vs Onset | 8.79 to 15.63 Hz | 49.565 | .00015 |
| Left | Post-dose vs Onset | 9.77 to 13.67 Hz | 18.562 | .01825 |

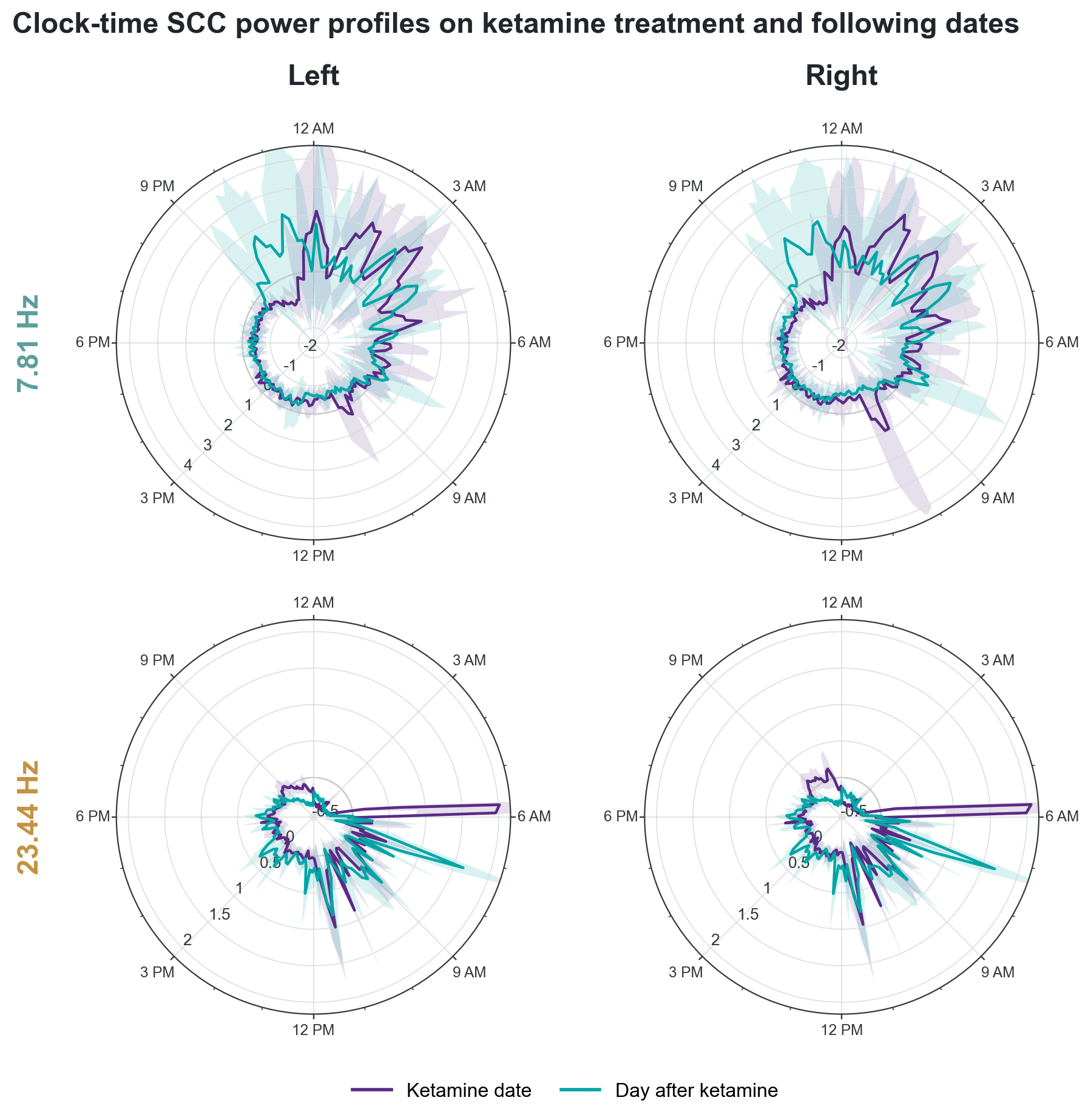

**Supplementary Fig. 1. Clock-time SCC power profiles on ketamine treatment and following dates.** Polar plots show mean within-day z-scored SCC power at 7.81 Hz (top) and 23.44 Hz (bottom) in the left and right hemispheres. Purple represents ketamine treatment dates and teal represents the following calendar dates; shaded regions indicate pointwise 95% confidence intervals across dates. Times are shown in local clock time.

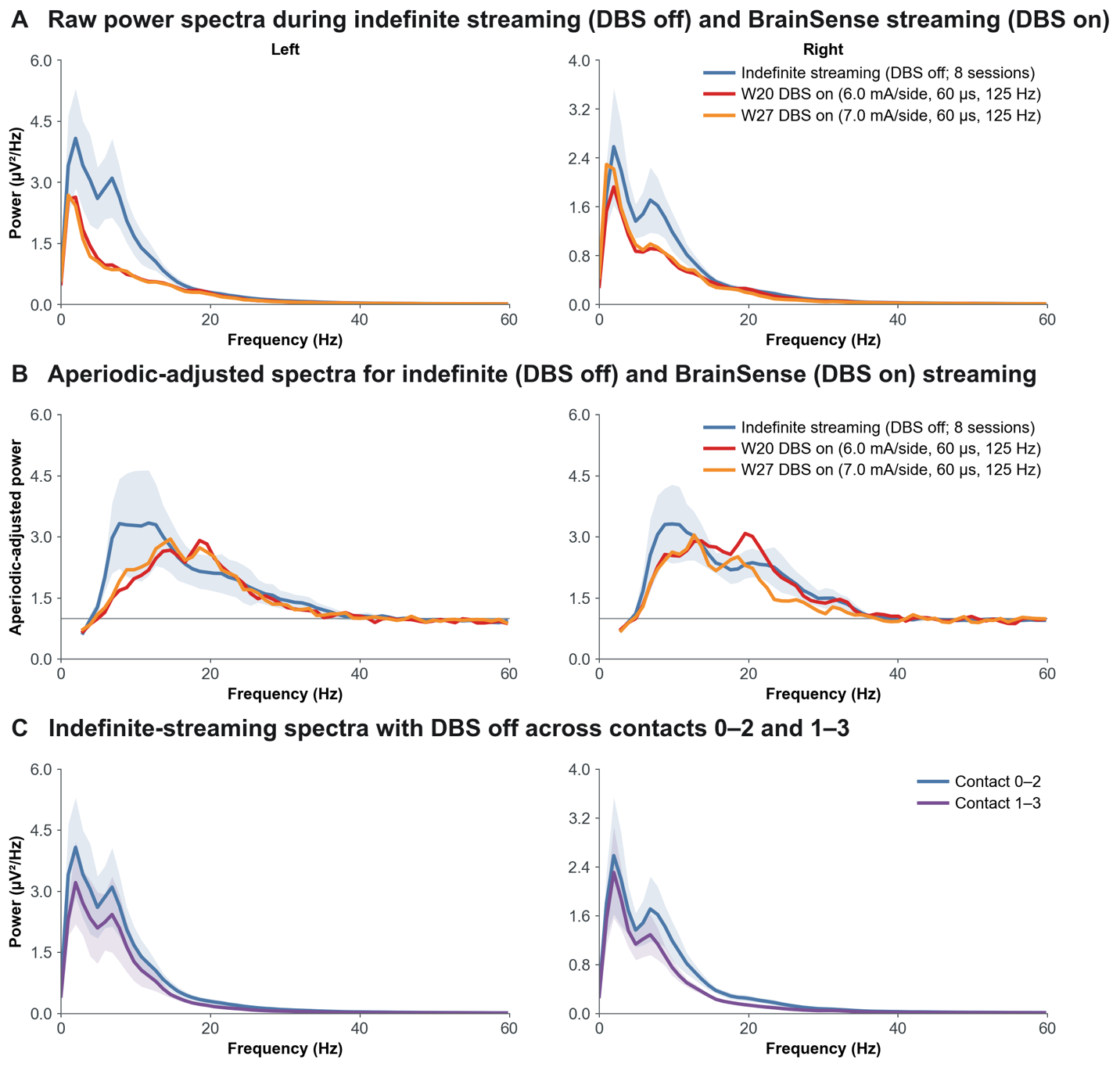

**Supplementary Fig. 2. Spectral profiles during DBS-off indefinite streaming and DBS-on BrainSense streaming.** (A) Raw power spectra from the left and right hemispheres during indefinite streaming with DBS off and BrainSense streaming with DBS on at Weeks 20 and 27. (B) Corresponding aperiodic-adjusted spectra. (C) DBS-off indefinite-streaming spectra recorded from contact pairs 0–2 and 1–3. For the indefinite-streaming data, lines show the mean and shaded regions indicate SEM across available recordings. These unmatched, nonrandomized recordings are shown descriptively. In the aperiodic-adjusted spectra, DBS-on recordings showed attenuated alpha-range peaks and a relative redistribution toward beta/high-beta activity.

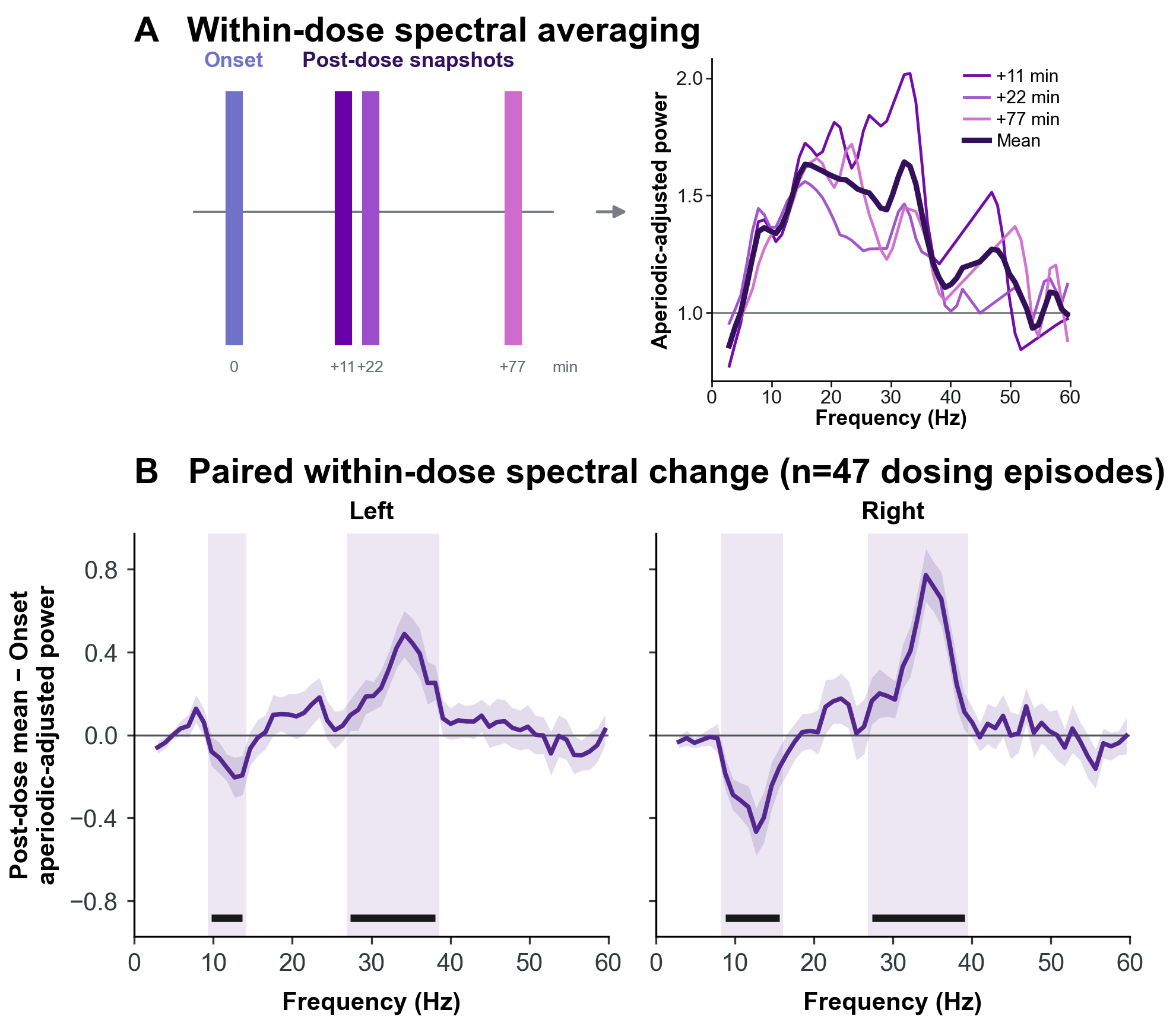

**Supplementary Fig. 3. Dosing-episode sensitivity analysis of ketamine-associated spectral changes.** (A) Medication Post-dose snapshots were averaged within each dosing episode and compared with the immediately preceding Medication Onset, yielding one paired difference per episode. (B) Mean Post-dose-minus-Onset aperiodic-adjusted power across 47 dosing episodes in the left and right SCC. Shading shows the pointwise 95% confidence interval. Purple shading and black bars mark significant frequency clusters from the sign-flip test with global maximum-cluster correction across hemispheres. Cluster ranges, masses, and corrected p values are reported in Supplementary Table 6.
